# Absolute and relative intensity physical activity of children with healthy and low levels of cardiorespiratory fitness

**DOI:** 10.1101/2025.06.25.25330324

**Authors:** Stuart J. Fairclough, Fabian Schwendinger, Sarah L. Taylor, Lynne M. Boddy, R. Glenn Weaver, Alex V. Rowlands

## Abstract

**Introduction:** Accelerometer-derived outcomes describing physical activity (PA) volume and intensity distribution relative to a person’s maximal capacity have provided insight into associations with health in adults. Little is known, however, about how the relative intensity of children’s PA volume and intensity distribution relates to health or fitness. To address this, we examined associations between children’s absolute and relative PA volume and intensity distribution with cardiorespiratory fitness (CRF), and differences in these PA outcomes for children stratified by CRF level.

**Materials and Methods:** In 9–10-year-old children (N=235) PA was assessed using wrist accelerometers for up to 7-days and CRF estimated from the 20-m multistage shuttle run test (20mSRT). Children were classified as Healthy or Low CRF. Absolute PA outcomes were PA volume (average acceleration; AvAcc_abs_) and intensity distribution (intensity gradient; IG_abs_). Equivalent relative PA outcomes were generated (AvAcc_rel_ and IG_rel_) using maximum acceleration values derived from the 20mSRT.

**Results:** Absolute, but not relative standardised PA outcomes were positively associated with CRF (AvAcc_abs_ Std*β*=0.21, *p*=0.02; IG_abs_ Std*β*=0.21, *p*=0.03). Absolute standardised PA outcomes were significantly higher among Healthy CRF children (AvAcc_abs_ Std*β*=0.40, *p*=0.007; IG_abs_ Std*β*=0.46, *p*=0.008), but there were no significant differences between Healthy and Low CRF groups for relative PA outcomes.

**Conclusions:** Children were similarly active relative to their physiological capacity, despite children with Healthy CRF being more active in absolute terms. Future studies should seek to better understand the influence of relative PA on CRF among diverse child populations who differ on a range of physical, physiological and demographic characteristics.

## Introduction

Accelerometer-assessed physical activity (PA) is commonly expressed in absolute intensity terms as energy expenditure-related time-use estimates (e.g., minutes of moderate-to-vigorous PA (MVPA)) (1). These are derived from population-specific intensity cut-points at the accelerometer data post-processing stage. Recent years have seen the emergence of directly assessed PA ‘cut-point-free’ average daily intensity (or volume) metrics such as average acceleration (2) and intensity profile metrics, such as the intensity gradient (2), which avoid the known limitations of population-specific absolute intensity cut-points (3). While it is standard practice to report time-use and cut-point-free accelerometer PA outcomes in absolute terms, this does not account for individual differences in physical capabilities, fitness, or health status. These factors are important because they influence the intensity at which PA is undertaken relative to an individual’s maximal capacity (4) which can be particularly significant when investigating PA associations with health indicators or evaluating PA intervention effectiveness. Thus, this ‘one size fits all’ approach may limit the utility of accelerometer data by introducing error into results derived from individuals whose fitness and health-related status (e.g., overweight/obese) vary (5). Cardiorespiratory fitness (CRF) in particular is an important consideration, as it has strong dose-response relationships with a range of health indicators through the life-course (6) and is a determinant of the intensity of PA that can be undertaken (1). Specifically, high absolute intensity of free-living PA is only possible in those with high CRF, while high PA intensity relative to CRF is generally attainable for all (7). Furthermore, over time PA of high relative intensity likely contributes to increased CRF and therefore health (1). However, while both absolute and relative intensity of PA are associated with CRF, the latter is rarely investigated.

Accelerometers measure the acceleration of the body part to which they are attached rather than the intensity-related physiological stress of PA. Nonetheless, studies have demonstrated that accelerometer PA outcomes can be adjusted to account for variations in CRF when individually calibrated against a physiological estimate of CRF such as oxygen uptake (1, 8) or heart rate (9, 10) and subjective indicators of relative intensity like rating of perceived exertion (RPE) (9, 11). Accelerometer studies that have used CRF to generate individually calibrated PA outcomes in child and adult populations have reported wide variations in PA time-use estimates when expressed in absolute terms and relative to CRF, and significant differences in absolute (e.g., 3 METs for MVPA) and relative PA intensity thresholds (1, 8, 10). These studies underscore the importance of estimating PA intensity relative to CRF when drawing conclusions about group differences in PA and associations with health indicators. However, integrating CRF assessments into PA study protocols, particularly in laboratory-settings is time and resource-intensive, placing additional burden on participants (12) and thus, is not routinely practiced.

Notwithstanding this, an approach for accelerometer-assessed relative PA intensity using a multistage maximal walking test was recently reported, whereby accelerations were recorded to estimate each participant’s maximum acceleration (i.e., maximal physical capacity) (12). This method can enable subsequent free-living accelerations to be expressed relative to maximum acceleration and thus participants’ relative PA intensity during free-living activities (12). Use of maximal field-based CRF assessments to estimate maximum accelerations for calculation of relative PA intensity therefore holds promise and overcomes the need for time- and resource-intensive laboratory testing. To our knowledge this approach has not been used with children.

The utility of accelerometer PA relative intensity outcomes has been demonstrated with cut-points in children (8) and adults (1) and with cut-point-free approaches in healthy adults (13) and clinical populations (7, 12). To date, no studies have investigated the relative intensity of children’s accelerometer-assessed PA using cut-point-free PA outcomes. Exploring relative intensity in children may provide a more nuanced approach to understanding their PA patterns and offer insights into associations with health, beyond that explained by ‘one size fits all’ absolute intensity PA outcomes. Thus, in this study we aimed to examine (1) associations between children’s absolute and relative PA outcomes and CRF and (2) differences in absolute and relative intensity PA outcomes for children stratified by CRF level.

## Materials and Methods

### Participants and settings

This is a secondary analysis of baseline data collected in phase 3 of the Active Schools: Skelmersdale PA pilot intervention study (14) (ClinicalTrials.gov registration: NCT03283904). Participant recruitment for study phases 2 (intervention design) and 3 (intervention evaluation) took place between 1^st^ May and 31^st^ October 2016. During this period 235 children aged 9-10-years old were recruited from seven primary schools situated in a low socio-economic status (SES) town in north-west England. Neighbourhood-level SES was calculated from home postcodes using the 2015 English Indices of Multiple Deprivation (EIMD) (15). EIMD rank scores ranging from 1 (most deprived area) to 32,844 (least deprived area) were generated for each child. Ethical approval was granted by Edge Hill University’s Faculty of Arts and Sciences Research Ethics Committee (reference #SPA-REC-2015–330) and written informed consent and assent were provided by the participants’ parents/carers, and the participants themselves, respectively. Collection of the baseline intervention data reported here took place in September 2017.

### Measures

#### Anthropometric measures

Height and body mass were measured using a portable stadiometer (Leicester Height Measure, Seca, Birmingham, UK) and calibrated scales (813 model, Seca), respectively. Body mass index (BMI) was calculated for each child and BMI z-scores were assigned (16) and International Obesity Task Force BMI cut-points applied to classify children as normal weight or overweight/obese (17). Sex-specific equations were used to predict age from peak height velocity (APHV), as a proxy measure of biological maturation (18). For all measurements children wore shorts and t-shirt with shoes removed.

#### Cardiorespiratory fitness

The 20-m multistage shuttle run test (20mSRT) provided an estimate of cardiorespiratory fitness (CRF). This test has been used extensively with similarly aged children to those in this study. The Leger et al. prediction equation (19) was applied to the running speed at the last completed lap to estimate peak oxygen uptake (V̇O_2peak_; ml‧kg^−1^‧min^−1^), which was used to represent CRF. Using a normative quintile-based framework, children were classified as having moderate to very high CRF (i.e., ‘Healthy’) or low to very low (i.e., ‘Low’) CRF levels based on the 40th centile cut-off for V̇O_2peak_ in European children (20).

#### Physical activity data generation

Children wore ActiGraph GT9X triaxial accelerometers (ActiGraph, Pensacola, FL, USA) on the non-dominant wrist for 24 hour·d^−1^ over 7 days with recording frequency set to 100 Hz. Data were downloaded using ActiLife version 6.11.9 (ActiGraph, Pensacola, FL, USA), and saved in raw format as GT3X files for processing using the GGIR R package v3.1-4 (21). Subsequent signal processing was then carried out in GGIR which included autocalibration using local gravity as a reference and detection of implausible values and of non-wear. Non-wear was imputed by default in GGIR whereby invalid data were imputed by the average at similar time points on other days of the week. Average waking (07:21 hours) and sleep onset times (22:17 hours) generated in the original study were used with the GGIR *qwindows* function to define the timing and duration of a valid ‘waking hours’ day. Wear time criteria were at least three valid days with ≥ 600 min·day^−1^ defined as a valid wear day. Accelerometer data were excluded from analyses if post-calibration error was > 10 m*g* (milli-gravitational units) and/or the wear time criteria were not achieved. The triaxial accelerometer signals were converted into one omnidirectional measure of acceleration (ENMO; i.e., the Euclidean norm of the three accelerometer axes with 1 *g* subtracted and negative values truncated to zero). Computed average valid day ENMO values expressed in m*g* were averaged over 5-s epochs and used to generate all subsequent PA outcomes.

*Absolute intensity physical activity outcomes (Figure 1)*.

**Figure 1.**
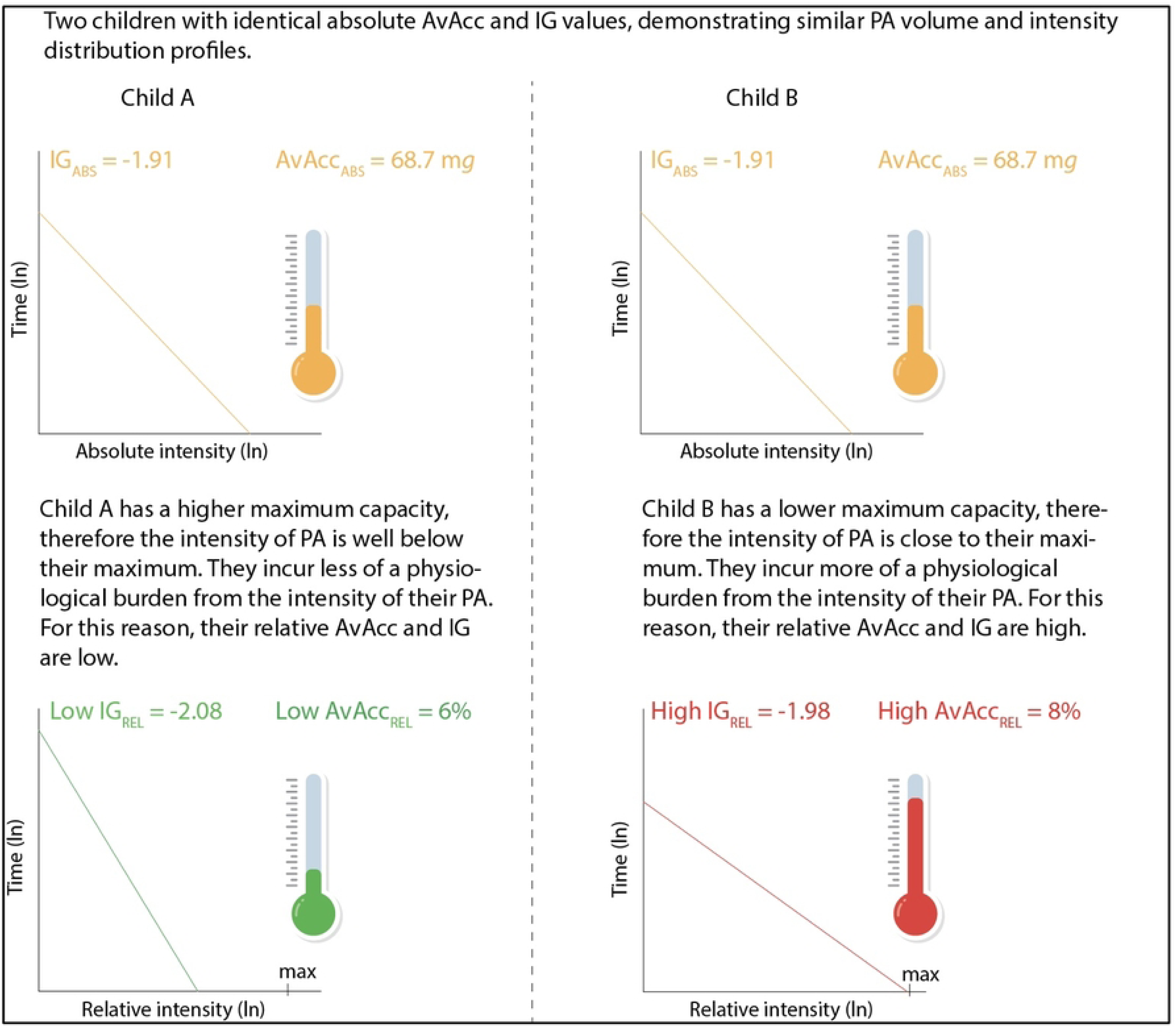
Visual interpretation of the absolute and relative PA outcomes. AvAcc: average acceleration; IG: intensity gradient; PA: physical activity; ln: natural log; m*g*: milligravitational units; Abs: absolute intensity; Rel: relative intensity; max: maximum.

*Absolute average acceleration (AvAcc_abs_)* represents the average intensity across the waking day (referred to as the ‘day’ from hereon); this is a proxy for PA volume. *Absolute intensity gradient (IG_abs_)* describes the PA intensity distribution across the day and reflects the negative curvilinear relationship between PA intensity and time accumulated at any given intensity (2). IG_abs_ values are always negative, with higher values indicating a greater proportion of PA is spent at higher intensities.

*Absolute MX metrics* (where X refers to an accumulated duration of time) represent the acceleration in m*g* above which the most active X minutes are accumulated (22). Absolute MX_abs_ metrics were computed for durations ranging from 2 hours (M120_abs_) to 1 min (M1_abs_).

*Relative intensity physical activity outcomes (Figure 1)*.

*Relative average acceleration (AvAcc_rel_)* expresses the PA volume relative to a child’s predicted maximum (aerobic) acceleration (i.e., AvAcc_abs_ / maximum acceleration *100) (13). Maximum acceleration was estimated by applying the children’s 20mSRT-derived VȮ_2peak_ values to the ENMO regression equation of Hildebrand et al. who reported a linear relationship between ENMO and VȮ_2peak_ (maximum acceleration (m*g*) = (V̇O_2_ (ml·kg^-1^·min^-1^) - 10.83) / 0.0356) (23).

*Relative intensity gradient (IG_rel_)* describes the relative intensity distribution of PA across the day, expressed in relation to a child’s maximum acceleration. IG_rel_ was derived from the epoch-level comma-separated values (.csv) files generated by GGIR using custom-built R script available at www.github.com/Maylor8/Relative-Intensity-Gradient (7).

*Relative MX metrics* (e.g., M120_rel_ to M1_rel_) were calculated using each child’s predicted maximum acceleration (i.e., MX_abs_ / maximum acceleration * 100) (13).

### Data analysis

Data analyses were performed in R (version 4.3.3) and R Studio (v2021.09.0). No data were available for three children who were absent on data collection days, and these cases were removed. Of the remaining 232 children, 173 achieved the three valid day accelerometer wear criteria, resulting in 25.4% data attrition. Visual inspection of the distribution and patterns of missing data and analysis of participant characteristics between children with and without PA outcome data confirmed there were no statistically significant differences in age, EIMD rank, sex, and BMI. We therefore proceeded with the assumption that the data were Missing at Random and used the *mice* package v. 3.17.0 (24) to perform multiple imputation by chained equations to replace missing values. Multiple imputation aims to minimise the impact of data attrition or non-response bias on data analysis by using available information about study participants to adjust parameter estimates, which can be subject to biases when data are missing (25). Multiple imputation can therefore approximate what results would look like with complete observations while allowing for representation of uncertainty in the results and maximising a dataset’s statistical power (26). A total of 25 multiply imputed data sets with 35 iterations were created as per recommendations to set *m* (i.e., the number of imputations) ≥ 100 times the highest fraction of missing information (25% fraction of missing information for PA outcomes (59/232)) (27). Intra-class correlations for the PA and CRF variables ranged from 0.01 to 0.06, which indicated low school-level influence on the PA outcomes and CRF. Results of subsequent analyses of each of the multiply imputed data sets were pooled as per Rubin’s Rules (28) (N=232).

Descriptive statistics were calculated and bivariate Pearson correlations computed between the absolute and relative PA outcomes and CRF. For study Aim 1, separate multiple regression models examined the associations of (a) absolute PA outcomes (AvAcc_abs_ and IG_abs_) with CRF, and (b) relative PA outcomes (AvAcc_rel_ and IG_rel_) with CRF. To address Aim 2, separate regression models investigated between-CRF group differences in absolute and relative PA outcomes. Sex, SES, and maturation were included as covariates. To enhance comparability all regression models expressed the PA outcomes as standardised (Std) estimates to reflect the different units of measurement for AvAcc_abs_, AvAcc_rel_, and IG_abs/rel_. These Std estimates were interpreted as small (≥0.1 to <0.3), medium (≥0.3 to <0.5), and large (≥0.5) effects (29).

Unstandardised regression model results are reported in Table S1 (Supporting Information File). MX_abs_ and MX_rel_ metrics were visualised descriptively using radar plots to describe patterning of absolute and relative intensity PA outcomes for Healthy and Low CRF groups. Absolute and relative intensity PA thresholds for moderate intensity PA (MPA) and vigorous intensity PA (VPA) were marked on the plots to aid interpretation of PA intensity. Absolute intensity thresholds were taken from widely used published cut-points of 201 m*g* (MPA) and 707 m*g* (VPA) (23). Relative intensity thresholds were calculated from acceleration values corresponding to CRF values of ≥46-63% V̇O_2peak_ (MPA) and ≥64% VȮ_2peak_ (VPA) respectively, as recommended in the American Heart Association’s Scientific Statement on youth CRF and health (6).

## Results

Descriptive characteristics of the sample are presented in Table 1 (N = 232). The children were 9.6 (0.3) years old and 50.9% were girls. The mean EMID rank of 6571.6 (7114.7) (out of 32844 ranked areas) indicated that area-level SES of the sample was relatively low. Mean VȮ_2peak_ was 47.6 (4.0) ml·kg^-1^·min^-1^ and 53.3% of the sample had Healthy CRF. Absolute daily PA volume (AvAcc_abs_) was 68.7 (22.0) m*g* and intensity distribution (IG_abs_) was −1.91 (0.18). AvAcc_rel_ and IG_rel_ were 7.0 (2.0)% and −2.15 (0.21), respectively.

**Table 1.** Descriptive characteristics of the participants (Mean (SD) or Frequency/%).

|  | Mean (SD)/Freq (%) |  |  |
| --- | --- | --- | --- |
|  | All<br>(N=232) | Healthy CRF<br>(N=122) | Low CRF<br>(N=110) |
| Age (y) | 9.6 (0.3) | 9.6 (0.3) | 9.6 (0.3) |
| Sex |  |  |  |
| Boys | 114 (49.1%) | 69 (56.6%) | 45 (40.9%) |
| Girls | 118 (50.9%) | 53 (43.4%) | 65 (59.1%) |
| EIMD rank | 6571.6 (7114.7) | 6094.0 (6175.2) | 6576.7 (7510.9) |
| Height (cm) | 137.2 (6.2) | 136.8 (6.1) | 138.2 (6.3) |
| Weight (kg) | 36.4 (18.2) | 36.1 (19.4) | 40.9 (18.8) |
| BMI (m<sup>2</sup>·kg<sup>-1</sup>) | 18.6 (3.3) | 18.5 (3.0) | 20.7 (3.7) |
| BMI z-score | 0.7 (1.2) | 0.7 (1.2) | 0.8 (1.2) |
| Weight status |  |  |  |
| Normal weight | 158 (68.1%) | 85 (69.7%) | 73 (66.4%) |
| Overweight/obese | 74 (31.9%) | 37 (30.3%) | 37 (33.6%) |
| APHV (y) | -2.7 (0.7) | -2.6 (0.7) | -2.7 (0.7) |
| 20mSRT laps | 30.8 (15.3) | 43.7 (11.8) | 26.0 (13.9) |
| VO<sub>2peak</sub> (ml·kg<sup>-1</sup>·min<sup>-1</sup>) | 47.6 (4.0) | 50.9 (3.1) | 46.4 (3.7) |
| Maximum acceleration (mg) | 1034.8 (111.8) | 1126.9 (85.8) | 997.5 (103.4) |
| Accelerometer wear (min·day<sup>-1</sup>) | 841.9 (68.8) | 839.9 (67.7) | 841.9 (70.0) |
| Physical activity outcomes |  |  |  |
| AvAcc<sub>abs</sub> (mg) | 68.7 (22.0) | 72.9 (20.3) | 67.2 (20.6) |
| IG<sub>abs</sub> | -1.91 (0.18) | -1.87 (0.17) | -1.92 (0.18) |
| AvAcc<sub>rel</sub> (%) | 7.0 (2.0) | 7.0 (2.0) | 7.0 (2.0) |
| IG<sub>rel</sub> | -2.15 (0.21) | -2.12 (0.20) | -2.16 (0.21) |

Footnotes. EIMD: English indices of multiple deprivation; BMI: body mass index; APHV: age from peak height velocity; 20mSRT: 20 m shuttle run test; AvAcc: average acceleration; IG: intensity gradient.

Correlations between the absolute and relative PA outcomes were stronger for daily PA volume (AvAcc_abs_ and AvAcc_rel_, *r*=0.81, 95%CI=0.80, 0.82) than the intensity distribution (IG_abs_ and IG_rel_, *r*=0.47, 95%CI=0.45, 0.49). Positive correlations of moderate magnitude were observed between AvAcc and IG, irrespective of whether they were expressed as absolute intensity (*r=*0.32, 95%CI=0.30, 0.34) or relative intensity (*r*=0.32, 95%CI=0.30, 0.35). Significant correlations were also evident between CRF and the PA outcomes, which were moderate for the absolute intensity outcomes (AvAcc_abs_: *r*=0.28, 95%CI=0.26, 0.30; IG_abs_: *r*=0.27, 95%CI=0.24, 0.29) and trivial-to-small for the relative intensity outcomes (AvAcc_rel_: *r*=0.04, 95%CI=0.02, 0.07; IG_rel_: *r*=0.17, 95%CI=0.14, 0.19).

<u>Aim 1.</u> Regression model results for study Aim 1 are summarised in Table 2. There were small positive associations with trivial to moderate effect sizes, between the absolute PA outcomes and CRF after adjustment for covariates (Model 1; AvAcc_abs_: Std*β*=0.21, 95%CI=0.04, 0.39; IG_abs_: Std*β*=0.21, 95%CI=0.02, 0.40). In contrast, there were no significant associations between either of the relative PA outcomes and CRF, although the standardised estimate for IG_rel_ (Std*β*=0.19, 95%CI=-0.01, 0.39) was similar to that for IG_abs_ and substantially higher than that for AvAcc_rel_ with effect sizes ranging from negligible to small (Model 2). Across both models the VIF values ranged from 1.00 to 4.27, indicating low risk of multicollinearity.

**Table 2.** Cross-sectional associations between absolute and relative physical activity outcomes and CRF.

| Term | Standardised estimates (95% CI) |  |
| --- | --- | --- |
|  | Model 1 | Model 2 |
| Intercept | -0.09 (-0.38, 0.18) | -0.10 (-0.39, 0.20) |
| AvAcc<sub>abs</sub> | **0.21 (0.04, 0.39)** | - |
| IG<sub>abs</sub> | **0.21 (0.02, 0.40)** | - |
| AvAcc<sub>rel</sub> | - | -0.03 (-0.22, 0.16) |
| IG<sub>rel</sub> | - | 0.19 (-0.01, 0.39) |
| Sex | 0.20 (-0.32, 0.72) | 0.19 (-0.35, 0.73) |
| SES | -0.06 (-0.19, 0.07) | -0.06 (-0.19, 0.08) |
| Maturation | 0.06 (-0.19, 0.32) | 0.04 (-0.23, 0.31) |

Footnotes. Bold text indicates statistically significant association with CRF; Sex, SES, Maturation included as covariates in both models; VIF = 1.02 to 4.24; AvAcc: average acceleration; IG: intensity gradient; SES: socioeconomic status; 95%CI: 95% confidence intervals.

<u>Aim 2</u>. AvAcc_abs_ and IG_abs_ were moderately higher among children with Healthy CRF compared to peers with Low CRF (AvAcc_abs_: Std*β*=0.40, 95%CI=0.11, 0.6; IG_abs_: Std*β*=0.46, 95%CI=0.12, 0.80). There was little evidence for between-CRF-group differences in AvAcc_rel_ (Std*β*=0.005, 95%CI=-0.31, 0.32) and IG_rel_ (Std*β*=0.30, 95%CI=-0.06, 0.66). For ease of interpretation the adjusted unstandardised mean differences between CRF groups are presented in Gardner-Altman estimation plots (Figures 2a-d) (30).

**Figure 2a-d.**
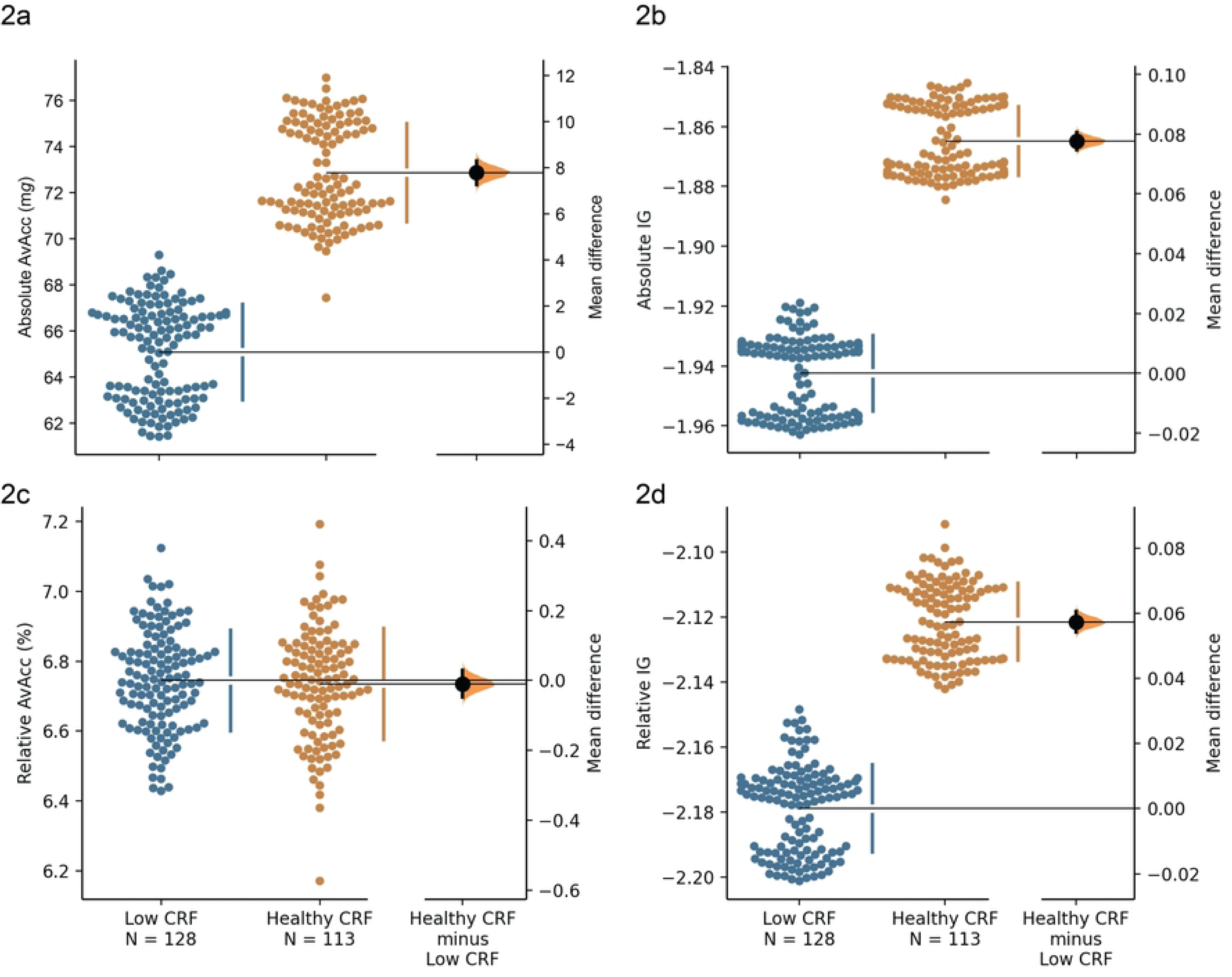
Gardner-Altman estimation plots displaying the adjusted mean differences between Low CRF and Healthy CRF groups. Both groups’ adjusted data are plotted on the left axes; the mean difference is plotted on a floating axes on the right as a bootstrap sampling distribution. The mean difference is depicted as a dot, with the 95% confidence interval is indicated by the ends of the vertical error bar. AvAcc: average acceleration; IG: intensity gradient; CRF: cardiorespiratory fitness; m*g*: milligravitational units.

PA patterning derived from absolute and relative intensity MX metrics in relation to CRF levels are described in Figures 3a and 3b (MX values are also presented in the Supporting Information File, Table S2). When absolute intensity thresholds were applied to MX metrics, children accumulated around 60 min·day^-1^ above moderate intensity irrespective of CRF level (Figure 3a). Healthy CRF children spent approximately 13 min·day^-1^ above the absolute intensity VPA threshold compared to ∼10 min·day^-1^ for children classified as having Low CRF. The Healthy CRF group was more active at higher intensities than Low CRF peers, particularly for time durations under 45 minutes. When intensity thresholds relative to the children’s maximum capacities were applied, the PA profile for Healthy and Low CRF groups were similar, with both spending a little over 20 min·day^-1^ and around 12-13 min·day^-1^ above relative MPA and VPA thresholds, respectively. Further, both groups were similarly active at all other MX time durations (Figure 3b).

**Figure 3a-b.**
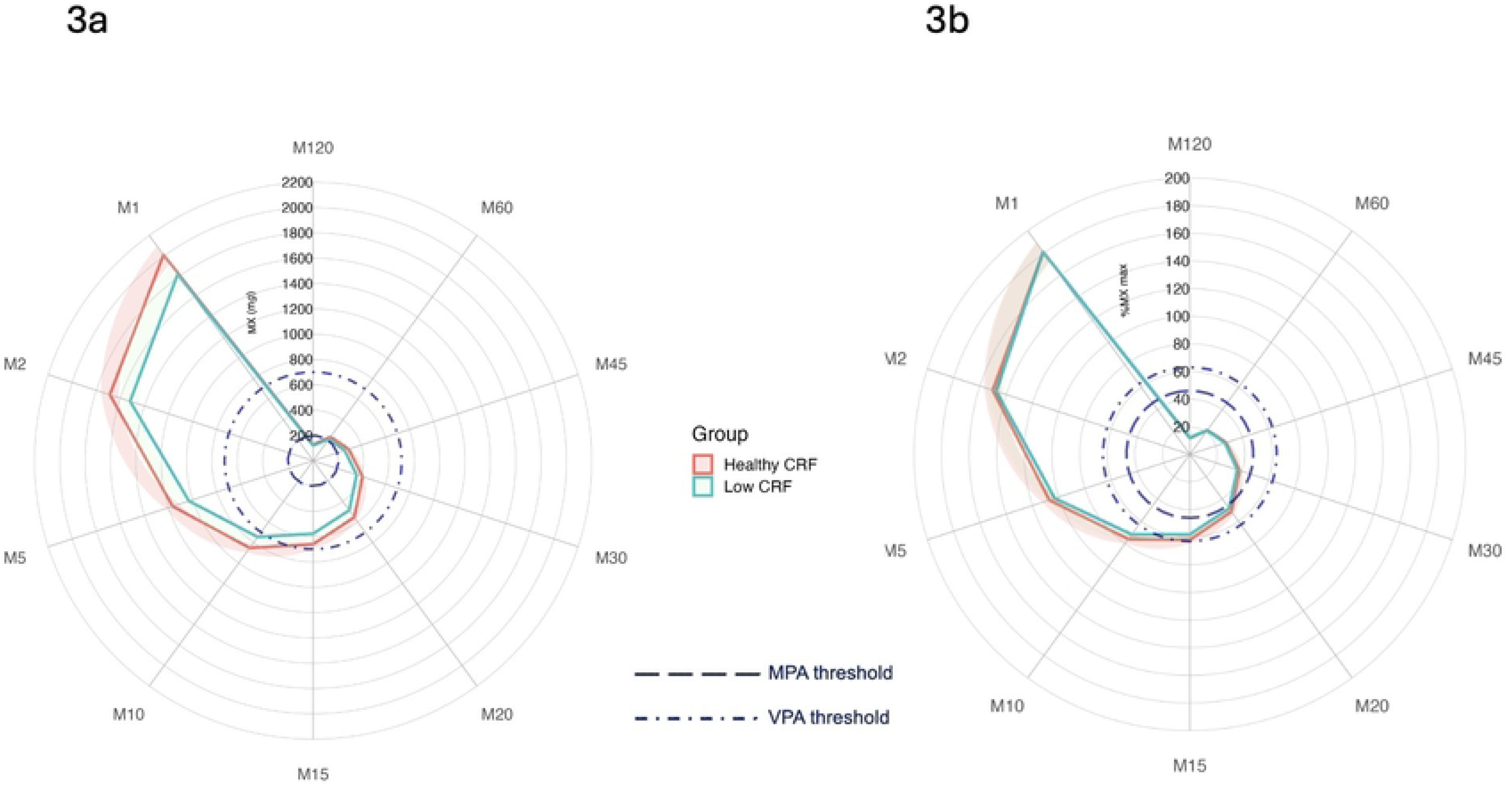
Radar plots illustrating absolute (3a) and relative (3b) MX metrics for the most active 120 to 1 min of the day for Healthy and Low CRF groups. m*g*: milligravitational units; %MX max: % of maximum acceleration.

## Discussion

This is the first study to report cut-point-free absolute and relative intensity PA outcomes in children. We found that absolute intensity PA outcomes were significantly and positively associated with CRF, and that IG_rel_ was more strongly associated with CRF than AvAcc_rel_. Consistent with these associations were the significant between-CRF group differences in absolute AvAcc and IG, but not in the relative intensity PA outcomes.

For Aim 1, absolute daily PA volume and intensity distribution across the day were similarly associated with CRF, with small-to-medium standardised effects. The association between absolute PA intensity and CRF indicates that the children with higher capacity generally used it during their daily activities. Further, the similarity of the associations for PA volume and intensity distribution suggests that the benefits of this higher capacity are evident in increased intensities of PA across the whole day and not just to higher intensity PA during discrete time periods. In a 2019 study with 10-year old children, IG_abs_ but not AvAcc_abs_ was independently associated with CRF (unstandardised *β*=13.79, 95%CI=9.34, 18.24; (31)), with the effect size greater than that observed in the current study (i.e., unstandardised *β*=4.71, 95%CI=0.23, 9.20) (31). The children in the current study were, however, more active than those in the 2019 sample with mean IG_abs_ values corresponding to ∼95^th^ centiles of reference values for northwest England children (32), compared to ∼85^th^ centile for the 2019 study (31). This is consistent with findings in healthy adults, where both AvAcc_abs_ and IG_abs_ outcomes were significantly associated with CRF, while only IG_abs_ had an independent association when the analysis was repeated in less active adults (a sample of patients with chronic heart failure) whose AvAcc_abs_ and IG_abs_ were 26.2% and 7.5% respectively, lower than more healthy and more active peers (33). In both low and high active populations, a greater breadth of the intensity distribution over the day (IG_abs_) is only possible when the person’s CRF is sufficient to enable engagement in higher intensities of PA. These higher intensities would also contribute to increasing CRF in both low and high active populations. Moreover, in more active populations AvAcc_abs_ may be more equally associated with CRF because the higher PA volume adds depth to the breadth of intensity experienced (i.e., especially at higher PA intensities), whereas in less active populations the breadth of the intensity distribution may dominate (33).

IG_rel_ had a similarly sized and positive association with CRF as IG_abs_, but with marginally wider confidence intervals that crossed zero. This suggests that regardless of fitness status, the children’s PA intensity distributions relative to their maximum capacities had a positive association with CRF (i.e., being active at a higher percentage of maximum capacity was associated with CRF). The relative intensity of the PA distribution can be interpreted using the ‘glass half-full’ analogy proposed by Orme et al. to illustrate different levels of PA engagement relative to maximum capacity (i.e., whereby the child’s maximum capacity is represented by the size of the glass and how full with water the glass is represents how much PA the child does) (12): The fitter children with higher maximum capacities and high IG_rel_ could be described as the ‘*can do*, *does do’* group (tall glass that is close to full), as their PA was high even relative to their greater maximum capacities. The less fit children, with high IG_rel_ could be described as the ‘*cannot do, does do’* group (short glass that is close to full), as their PA was also high relative to their capacities which were lower than fitter peers. In contrast, high-fit children with low IG_rel_ would be ‘*can do, does not do*’ (tall glass, less water) and low-fit children with low IG_rel_ would be ‘*cannot do, does not do*’ (short glass, not much water). How full the glass is (IG_rel_) irrespective of glass size was associated with CRF, though the effect size was small and uncertain. Because of this, it is probable that factors other than capacity were impacting on the intensity of the children’s PA. These may have included the genetic component of CRF as well as psychological, interpersonal, environmental, and demographic factors which in combination can favourably enable and predispose children to be active (34).

The association between AvAcc_rel_ and CRF was trivial, indicating that the relative PA volume was unrelated to the children’s maximum capacities. AvAcc_rel_ is reflective of activities of children’s daily living that are often disproportionately skewed towards the lower end of the relative intensity spectrum (e.g., classroom lessons, sedentary screen time, household chores, etc. (35)) which require relatively low physiological effort. When the large time contribution that these activities make across the day and therefore to the daily volume of PA is considered, the negligible association of AvAcc_rel_ with CRF was unsurprising.

Aim 2’s findings are consistent with the Aim 1 results, demonstrating that absolute but not relative PA volume and intensity distribution were significantly higher among the Healthy CRF group. This indicates that the lower PA intensities engaged in by children with Low CRF were proportionate to their reduced CRF (i.e., they were just as active relative to their maximum capacities as peers with Healthy CRF). This is consistent with the observation in adults that the absolute but not relative intensity of PA participated in declines as people age (13, 36, 37). Descriptive analysis using the MX metrics and radar plots illustrates this, with the higher absolute intensities of the Healthy CRF group’s most active 45 minutes to 1 minutes of the day clearly evident in Figure 3a and Table S2 (Supporting Information File), while the relative intensity of the two groups were very similar (Figure 3b and Table S2 Supporting Information File). For both CRF groups, PA time above moderate intensity was ∼ 40 min·day^-1^ lower when the relative intensity MPA threshold (6) was applied compared to when the frequently used absolute intensity MPA threshold for wrist-worn ActiGraph accelerometers was used (23). This aligns with findings from adult studies reporting higher MVPA derived from absolute versus relative intensity cut-points (9). Furthermore, it raises the question of whether the Hildebrand et al. (23) absolute MPA threshold adequately reflects estimates of children’s health-enhancing MPA. This query is all the more pertinent for children with high CRF, which is a point that has been posed previously in relation to other child and adult absolute intensity accelerometer cut-points for MPA/MVPA (1, 8, 10). In contrast, time above the absolute and relative VPA intensity thresholds were analogous (i.e., CRF group differences of around 3 min·day^-1^ (absolute intensity threshold) and 1 min·day^-1^ (relative intensity threshold)). This indicates that the absolute and relative VPA intensity thresholds were more comparable for this sample than the MPA thresholds. Moreover, the mean between-CRF group difference in MX_rel_ values across the M120 to M1 durations was 2.7%, compared to a 10.5% difference in MX_abs_ (Supplementary Table 2). These similarities in the relative intensity of PA volume between the Healthy and Low CRF groups suggest that physiological capacity may, in part, determine PA level. However, it is also well established that children’s PA determinants are multidimensional (34). Psychosocial correlates of children’s PA (38) in particular, such as affective valence and enjoyment influence PA behaviours especially at higher intensities, and for some children could play just as influential a role in their engagement than physiological capacity and perceived exertion (39).

This is the first study to report cut-point-free relative intensity PA outcomes in children. The calculation of AvAcc_rel_ and IG_rel_ as contemporary relative intensity PA outcomes derived from estimates of children’s CRF and maximum accelerations is an important strength which enabled the nuanced influences of absolute and relative PA on CRF to be examined. We used the 20mSRT as an accessible applied field-based CRF assessment that is commonly used in schools and other settings, which enhanced the ecological validity of the study. Further, use of multiple imputation maintained the integrity of statistical relationships in the data and reduced the risk of sampling and selection biases (25) due to missing data from accelerometer non-wear. Our findings are though caveated by some limitations. The cross-sectional study design limits conclusions about causality, and the results may not be generalisable to other geographic locations. Moreover, as our sample was relatively more active than a reference population of northwest England 10-year olds (32), caution is advised when interpreting the absolute PA outcomes. The children’s V̇O_2peak_ values were estimated from 20mSRT performance data and although this well-established test has moderate criterion validity in healthy children (40) some error was likely present in the V̇O_2peak_ values. The 20mSRT was though, the most pragmatic approach available for this study. Additionally, it enabled novel use of the Hildebrand et al. VȮ_2_ regression equation (23) to calculate maximum acceleration values (i.e., maximum PA capacity) and therefore the relative PA outcomes. Lastly, the significance of the genetic component of CRF is acknowledged as an example of an unmeasured confounder that influences capacity and is therefore a further limitation in our study.

In this study of cut-point-free absolute and relative intensity PA outcomes in children, absolute intensity PA outcomes were significantly and positively associated with CRF and were significantly higher in Healthy CRF children than those with Low CRF. Associations between relative intensity PA outcomes and CRF were weaker and less consistent, although IG_rel_ was more strongly associated with CRF than AvAcc_rel_. The lack of a difference in relative intensity PA outcomes between CRF groups indicates that children were similarly active relative to their physiological capacity. Quantifying both the absolute and relative intensity of accelerometer-assessed PA outcomes provides greater insight than either alone. The non-significant associations between the relative intensity PA outcomes and CRF suggests that this sample of highly active children may have been enabled and predisposed to engage in PA by factors other than, or as well as, their maximum aerobic capacities for PA. Future studies should build on these findings to better understand the influence of relative PA on CRF among diverse child populations who differ on a range of physical, physiological and demographic characteristics.

## Data Availability

The data that support the findings of this study are openly available from the Open Science Framework https://osf.io/wnmvs/files/osfstorage

## Acknowledgements

We acknowledge the help and engagement of the children and teachers in the participating schools.

## Author Contributions

SJF, AVR, LMB FS, SLT designed the study. SJF, SLT contributed to data collection. SJF, SLT analysed the data. SJF, AVR, LMB FS, SLT, RGW drafted the manuscript. All authors contributed to writing, editing, reviewing and approved the final manuscript. Funding for the original study was acquired by SJF.

## Ethics approval statement

Ethical approval was granted by Edge Hill University’s Research Ethics Committee (# SPA-REC-2015–330).

## Supporting information captions

Table S1. Cross-sectional associations between absolute and relative physical activity outcomes and CRF

Table S2. Mean (SD) absolute and relative MX metrics by CRF group

